# Cohort Profile: A national prospective cohort study of SARS-CoV-2 pandemic outcomes in the U.S. - The CHASING COVID Cohort Study

**DOI:** 10.1101/2020.04.28.20080630

**Authors:** McKaylee M. Robertson, Sarah Gorrell Kulkarni, Amanda Berry, Chloe Mirzayi, Madhura Rane, Mindy Chang, Shivani Kochhar, William You, Andrew Maroko, Rebecca Zimba, Drew Westmoreland, Christian Grov, Angela Parcesepe, Levi Waldron, Denis Nash, for the CHASING COVID Cohort Study

## Abstract

**Purpose:** The CHASING COVID Cohort study is a U.S.-based prospective cohort study launched during the upswing of the U.S. COVID-19 epidemic. The objectives are to: 1) estimate and evaluate determinants of the cumulative incidence of SARS-CoV-2 infection, disease, and deaths; 2) assess the impact of the pandemic on psychosocial and economic outcomes; and 3) assess the uptake of pandemic mitigation strategies.

**Participants:** We began enrolling participants March 28, 2020 using internet-based strategies. Adults ≥18 years residing anywhere in the U.S. or U.S. territories were eligible. 6,753 people are enrolled in the cohort, including participants from all 50 U.S. states, the District of Columbia, Puerto Rico, and Guam. Participants are contacted regularly to complete study assessments, including interviews and specimen collection.

**Findings to date:** Of 4,247 participants who provided a specimen for baseline serologic testing, 135 were seropositive by screening antibody testing (3.2%, 95% CI 2.7%-3.5%) and 90 were seropositive by confirmatory antibody testing (2.1%, 95% CI 1.7%-2.6%). Cohort data have been used to assess the role of household crowding and the presence of children in the household as potential risk factors for severe COVID-19 early in the U.S. pandemic; to describe the prevalence of anxiety symptoms and its relationship to COVID-19 outcomes and other potential stressors; and to identify preferences for SARS-CoV-2 diagnostic testing when community transmission is on the rise via a discrete choice experiment.

**Future plans:** The CHASING COVID Cohort Study has outlined a research agenda that involves ongoing monitoring of the cumulative incidence and determinants of SARS-CoV-2 outcomes, mental health outcomes and economic outcomes. Additional priorities include COVID-19 vaccine hesitancy, uptake and effectiveness; incidence, prevalence and correlates of long-haul COVID-19; and the extent and duration of the protective effect of SARS-CoV-2 antibodies.

## INTRODUCTION

The Coronavirus Disease 2019 (COVID-19) pandemic has dramatically transformed life across the entire United States, resulting in medical and economic challenges and threats for individuals, households and communities. The earliest research efforts have focused on understanding the clinical course of COVID-19 and the most effective ways of treating people with severe symptoms or illness. As the pandemic progresses, however, we must also investigate COVID-19’s evolving epidemiology and the uptake/impact of non-pharmaceutical interventions (NPIs)^1^, such as physical distancing, health messaging, and testing on the cumulative incidence of SARS-CoV-2. Researchers and public health practitioners have called for cohort studies to describe the community attack rate, as well as how attack rates are influenced by different approaches to NPI implementation.^2^ There is also a need to characterize both the direct and indirect effects of the SARS-CoV-2 pandemic on mental health and economic outcomes. Internet-based strategies, which facilitate rapid recruitment of large and diverse samples, can be leveraged to understand and inform this swiftly changing and protracted public health crisis.^3,4^

In response to the COVID-19 pandemic the CUNY Institute for Implementation Science in Population Health (ISPH) launched the prospective Communities, Households and SARS-CoV-2 Epidemiology (CHASING) COVID Cohort study on March 28, 2020. We sought to prospectively recruit a geographically and socio-demographically diverse cohort of adults (18 years or older) in the United States (U.S.) and U.S. territories in order to contribute to our understanding of the spread and impact of the SARS-CoV-2 pandemic.

## COHORT DESCRIPTION

### Objectives and study design

Key objectives of the cohort study are to: 1. Estimate and evaluate determinants of the cumulative incidence of SARS-CoV-2 infection, disease, and deaths; and 2. Assess the pandemic’s impact on psychosocial and economic outcomes (mental health, unemployment, food security); and 3. Assess the uptake of pandemic mitigation strategies (non-pharmaceutical interventions, testing, contact tracing, isolation/quarantine, vaccination). The study design is shown in Figure 1. Study visits (completion of questionnaires online and designated by V_x_) are completed every 1-3 months following cohort screening and enrollment, and will continue through December 2021, for a maximum of 21 months follow-up. Initial specimen collection (S_1_) occurred during April-June 2020 and the second specimen (S_2_) will be collected in November 2020-January 2021. Additional specimen collection may occur in 2021, depending on epidemic activity and availability of funding.

**Figure 1:**
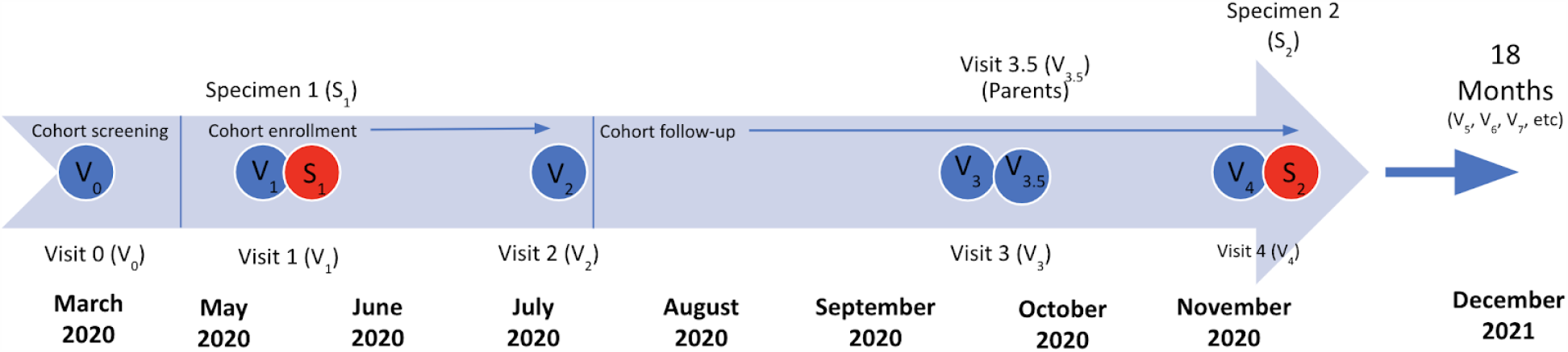
Design for the CHASING COVID Cohort Study.

### Cohort eligibility

Eligibility was determined during cohort screening visits and cohort enrollment visits (Figure 2). To be eligible for inclusion in the cohort, individuals had to: 1) reside in the U.S. or a U.S. territory; 2) be age 18 years or older; 3) provide valid email address; and 4) demonstrate early engagement in longitudinal study activities, including: a) completion of V_1_ (which provided the opportunity to consent for serologic testing); and b) completion of at least one additional screening visit in addition to V_1_ (i.e., V_0_ or V_2_) or provision of a baseline specimen for serologic testing (S_1_).

**Figure 2.**
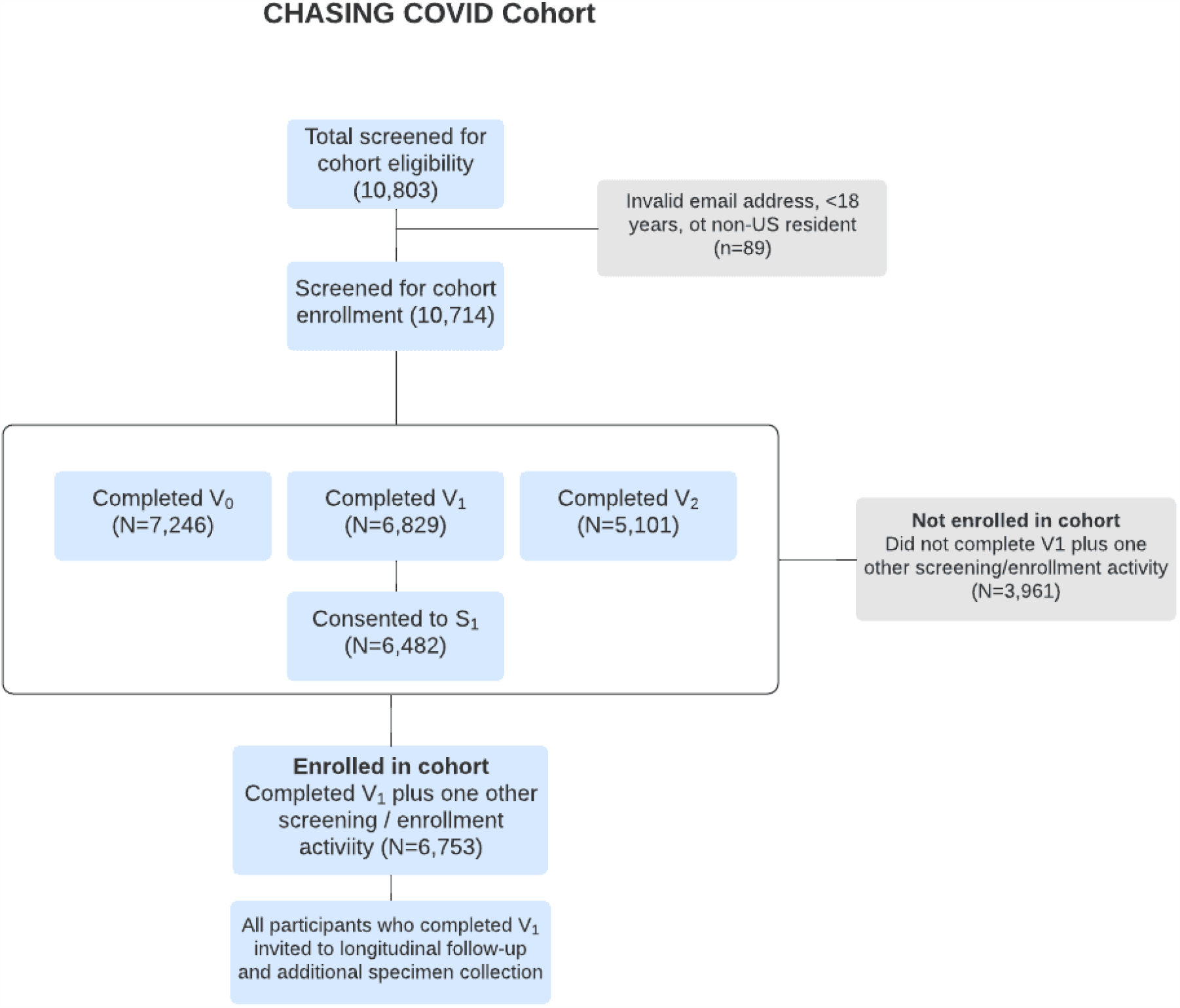
Enrollment and follow-up in the CHASING COVID Cohort.

### Cohort screening and enrollment

Cohort screening and enrollment began on March 28, 2020, at which point there were 122,000 documented COVID-19 cases and 2,200 COVID-19 deaths reported in the U.S..^5^4 Enrollment ended on August 21, 2020, when there were 783,000 persons diagnosed with SARS-CoV-2, including 42,000 deaths in the U.S..^5^

We used internet-based strategies that are effective for recruiting and following large and geographically diverse online cohorts, including at-home specimen collection.^3,4,6^ Persons aged 18 years and above who resided in the U.S. or U.S. territories were eligible to join the study. Study participants were recruited via ads on social media platforms (e.g., Facebook, Instagram, and Scruff), Qualtrics Panel, or via referral to the study (anyone with knowledge of the study was allowed to invite others to participate). By relying on personal networks of participants through referrals, we aimed to bolster recruitment of persons >59 years of age, who were important to have represented in the cohort because of their risk, but may not be as active on social media as younger persons. Facebook and Instagram advertisements were developed in English and Spanish and were geographically targeted to people currently residing in the U.S. and U.S. territories who were 18 or older.

Study staff systematically monitored cohort demographics and proactively adjusted advertisement strategies as needed to balance geographic and sociodemographic characteristics of respondents. For example, strategies could shift to prioritize recruitment of older persons if that demographic was poorly represented.

Interested participants were directed to a pre-enrollment survey (hosted by Qualtrics) to be completed in their web browser on a computer or on a mobile device.^7^ A consent form described the study, plans for follow-up assessments, and future study opportunities, including the possibility to receive SARS-CoV-2 serologic testing as part of the study. The consent form also described the incentive schedule: a drawing for $100 for a pre-enrollment survey (V_0_) (with 20 winners) and gift cards ranging from $5-30 for all participants for completion of subsequent surveys and antibody testing.

### Study measurements

Measures included on the screening, enrollment and follow-up study questionnaires were derived from previously published research (e.g., *Together 5000*^*3*^, BRFSS, and H1N1 influenza studies^8,9^) and from other researchers who had developed surveys for understanding COVID-19 (e.g., Canadian Institutes of Health Research^7^ and Food Access and Food Security during COVID-19^10^). Measures were also developed *de novo* in response to the novel pandemic. The follow-up assessments gather data on symptoms, testing, hospitalizations and other time-varying factors (see Table 1 for key measurement realms). All study questionnaires are available on CHASING COVID Cohort Study webpage (https://cunyisph.org/chasing-covid/).

**Table 1:** Realms of Measurement in the CHASING COVID Cohort.

|  | Measure | Cohort screening (V0) | Cohort enrollment (V1) | Follow-up (V2) | Follow-up (V3) | Parents |
| --- | --- | --- | --- | --- | --- | --- |
| Individual and household characteristics | Sociodemographics (various measures) | ✓ | ✓ | ✓ |  | ✓ |
|  | Household size and age distribution | ✓ |  |  |  | ✓ |
|  | Educational/childcare experience of household children |  |  |  | ✓ | ✓ |
|  | Height and weight |  |  | ✓ |  |  |
|  | Employment status | ✓ | ✓ | ✓ | ✓ | ✓ |
|  | Food and housing security |  | ✓ | ✓ |  |  |
| COVID-19 risk factors | Pre-existing/co-morbid conditions | ✓ |  |  |  |  |
|  | Smoking and vaping behaviors | ✓ |  |  |  | ✓ |
|  | Essential employee status* | ✓ | ✓ | ✓ | ✓ |  |
|  | NPI uptake and other precautions | ✓ | ✓ | ✓ | ✓ |  |
|  | Influence of social media on COVID knowledge and beliefs | ✓ | ✓ |  |  |  |
|  | Understanding of epidemiological concepts (antibodies, herd immunity) |  |  |  | ✓ |  |
| Health care-related | Health care access and insurance status | ✓ | ✓ | ✓ | ✓ |  |
|  | Sexual and reproductive health |  |  |  | ✓ |  |
| COVID-19-related outcomes | Recent COVID-like symptoms | ✓ | ✓ | ✓ | ✓ |  |
|  | Medical care for COVID-like symptoms | ✓ | ✓ | ✓ | ✓ |  |
|  | Recent SARS/COV2 testing** and diagnosis | ✓ | ✓ | ✓ | ✓ |  |
|  | Contact tracing***, quarantine, and isolation |  |  | ✓ | ✓ |  |
|  | Serology (SARS/COV2 IgM and IgG) |  | ✓ |  |  |  |
|  | Vaccine uptake |  |  | ✓ | ✓ |  |
| Mental and physical health | Anxiety (GAD-7) and risk perceptions | ✓ | ✓ | ✓ | ✓ | ✓ |
|  | Depression (PHQ8) |  | ✓ | ✓ |  | ✓ |
|  | PTSD (PC-PTSD-5) |  |  |  |  | ✓ |
|  | Resilience, stress, and coping |  |  |  |  | ✓ |
|  | Social support |  | ✓ | ✓ | ✓ | ✓ |
|  | IPV |  | ✓ | ✓ | ✓ |  |
|  | Drug use |  | ✓ |  |  | ✓ |
|  | Alcohol use (AUDIT) |  | ✓ |  |  | ✓ |
|  | Physical Activity |  |  |  | ✓ |  |

\*front-line health care workers, hospital staff, first responders, grocery clerks, delivery workers, educators (V3 only)
\*\* V3 covers testing experience and expanded testing questions
\*\*\* V3 covers informal contact tracing as well as onward transmission

### Daily symptom tracking

Study questionnaires are supplemented by voluntary daily symptom tracking via an innovative COVID-19 symptom tracker^11^ that we have deployed in our cohort. The Coronavirus Pandemic Epidemiology (COPE) consortium ^11^ has developed the COVID Symptom Tracker app, which enables individuals to self-report information on COVID-19 via daily prompts for recent symptoms, health care visits, and COVID-19 testing results. The CHASING COVID Cohort Study joined the COPE consortium on April 6th, allowing our cohort members who use the app to affiliate with our cohort and consent to have their their app responses linked with the CHASING COVID Cohort Study data.

### Specimen collection for serologic testing

At the V_1_ assessment (end of April, 2020 through July, 2020) and the V_4_ assessment (beginning November, 2020), participants were asked to self-collect a specimen at home for serologic testing. Participants are mailed dried blood spot (DBS) specimen collection kits. To facilitate self-sampling procedures, all participants are provided printed instructions and a QR code to view a video demonstrating procedures for DBS collection, and instructions to contact the study team, if they have questions.^12^ DBS cards are returned via the U.S. Postal Service (self-addressed, stamped envelope containing EBF Foil biohazard bag™) to the study laboratory (Molecular Testing Labs).^13^ Participants receive $20 upon receipt of a valid specimen by the study laboratory.

### Serologic testing

All DBS specimens are screened for SARS-CoV-2 antibodies using the Biorad total antibody test, and specimens that are reactive are tested further using the Euroimmun IgG test. Once tested, DBS specimens are banked at -80°C for future SARS-CoV-2 studies.

### Data management and analysis

All data were imported and cleaned in R and SAS (V9.4). Data were geocoded based on a self-reported ZIP code. Maps were created in ArcGIS 10.7. Data from V_1_ were used to compile summary statistics on baseline characteristics.

### Ethical Approval

The study protocol was approved by the Institutional Review Board at the City University of New York (CUNY) Graduate School for Public Health and Health Policy.

### Cohort Eligibility

Of the 10,803 individuals who completed at least one study screening or enrollment visit, 10,714 were age 18 or older, U.S. residents, and provided a valid email address for study follow-up. Of those, 7,246 completed V_0_, 6,829 completed V_1_, 5,101 completed V_2_, 6,482 consented to provide a baseline DBS specimen; 6,753 met final study eligibility criteria of completing 2 of 3 screening/enrollment visits or consenting to provide a specimen as part of V_1_, and were considered enrolled in the cohort (Figure 2). Participants who only completed V_1_ are routinely invited to complete additional study assessments and specimen collection.

## FINDINGS TO DATE

### Cohort Characteristics

The cohort includes 6,753 participants from all 50 states, the District of Columbia, Puerto Rico and Guam (Figure 3). At V_1_, the median age of participants was 37 years (interquartile range: 29, 51); 73% were aged 18-49 (including 5% <21 years (N=370)), 12% were aged 50-59, and 15% were aged 60 years or older (Table 2). Just under half (45%) were male, 20% were Hispanic, 13% black non-Hispanic, 7% Asian or Pacific Islander, and 57% white non-Hispanic. A majority were currently employed (63%), 10% were retired, 13% were out of work, and 9% were students.

**TABLE 2:**
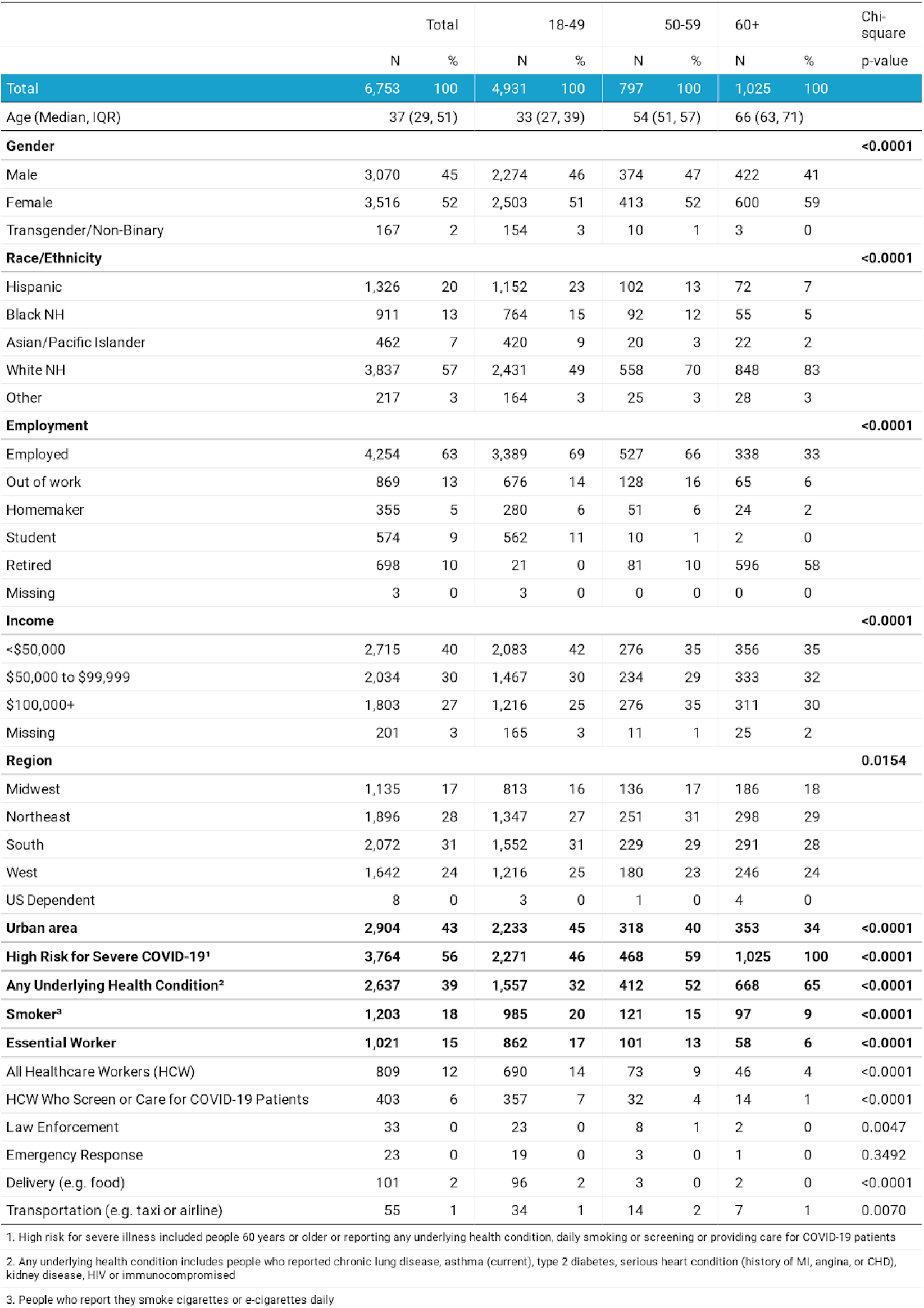
Characteristics of CHASING COVID Cohort.

**Figure 3.**
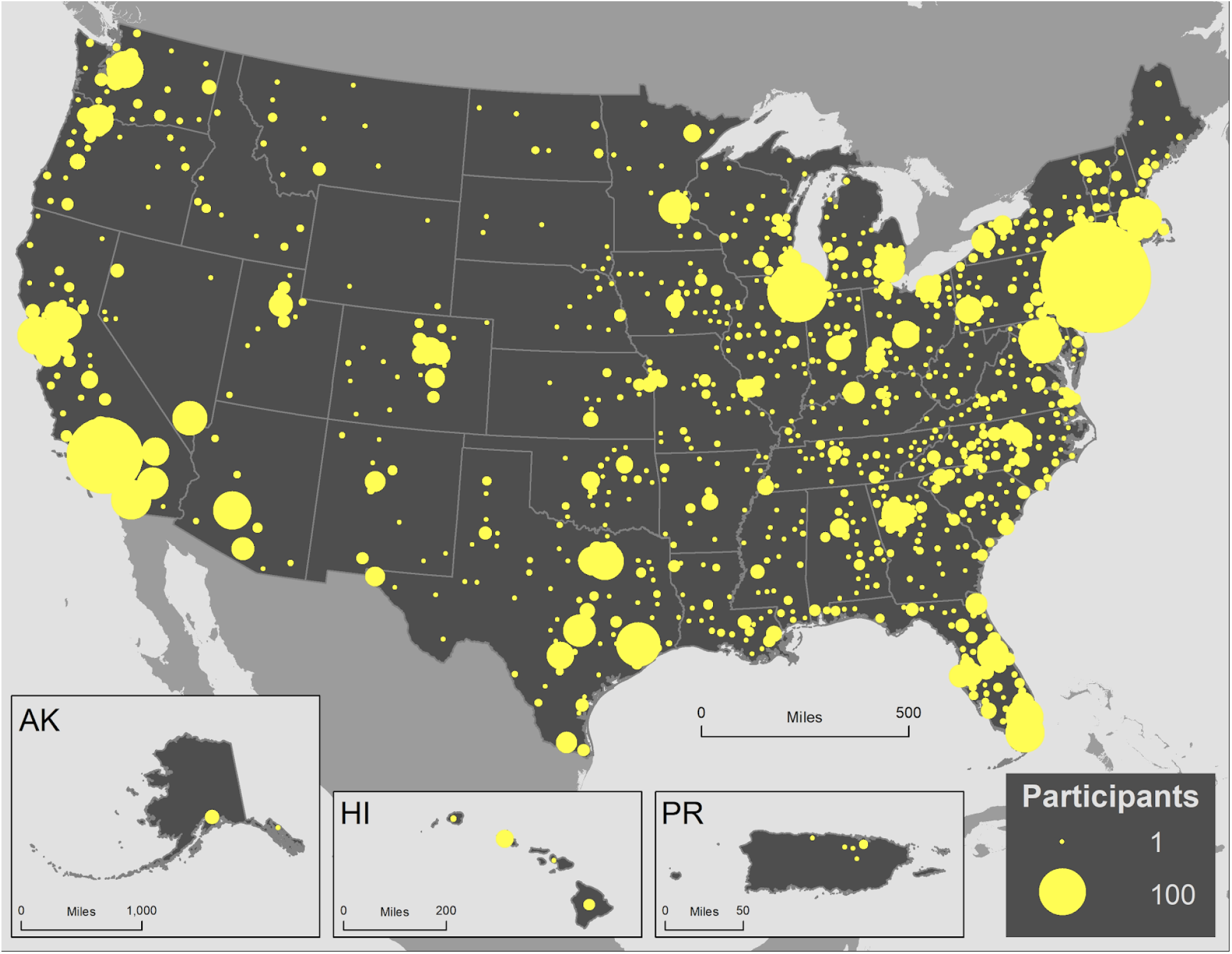
Geographic distribution of CHASING COVID Cohort participants by county of residence, N = 6,753.

More than half (56%) were considered to be at increased risk for severe COVID-19 disease should they become infected with SARS-CoV-2, on the basis of age (60+), presence of an underlying health condition (chronic lung disease, asthma [current], type 2 diabetes, serious heart condition, kidney disease, or an immunocompromised status), or daily smoking (Table 2). The proportion of persons with an underlying health condition increased with age category (32% among 18-49 year olds and 65% among 60+), and the proportion of daily smokers decreased with increasing age category (20% and 9%, respectively).

15% of participants reported being an essential worker (Table 2). By employment category, 12% were healthcare workers (half of whom reported screening or caring for COVID-19 patients), 1.5% worked in delivery services (e.g., food) and less than 1% worked in transportation (e.g., taxis). The proportion of persons employed in essential work decreased with increasing category of age.

### NPI / Physical Distancing Behaviors Stratified by Age Categories

A high proportion of participants reported avoiding large groups with >20 people in the prior two weeks and avoiding handshakes or hugs (89% and 88%, respectively) (Table 3). One-quarter (25%) reported working from home. A majority reported wearing gloves (58%) and masks (93%), and these proportions significantly increased with age (57% of 18-49 year olds wore gloves versus 60% of 60+, and 92% of 18-49 year olds wore masks versus 95% of 60+, p for chi-square: <0.001 for each comparison). Almost one in three (29%) participants reported stockpiling personal protective equipment and 39% reported stockpiling food. The proportion of participants who reported stockpiling decreased significantly with increasing age categories (31% of 18-49 year olds stockpiled PPE versus 24% of 60+, and 42% of 18-49 year olds stockpiled food versus 29% of 60+, p<0.001 for each comparison).

**Table 3:** Behaviors and Symptoms by Age Category of CHASING COVID Cohort.

|  | Total |  | 18-49 |  | 50-59 |  | 60+ |  | Chi-Square |
| --- | --- | --- | --- | --- | --- | --- | --- | --- | --- |
|  | N | % | N | % | N | % | N | % | P-value |
| Total | 6,753 | 100 | 4,931 | 100 | 797 | 100 | 1,025 | 100 |  |
| **Behaviors in the past month** |  |  |  |  |  |  |  |  |  |
| Avoided Groups >20 | 6,025 | 89 | 4,337 | 88 | 734 | 92.1 | 954 | 93 | <0.0001 |
| Avoided Handshakes | 5,969 | 88 | 4,272 | 87 | 746 | 93.6 | 951 | 93 | <0.0001 |
| Bought Firearm | 382 | 6 | 357 | 7 | 15 | 1.88 | 10 | 1 | <0.0001 |
| Worked from Home | 1,673 | 25 | 1,413 | 29 | 158 | 19.82 | 102 | 10 | <0.0001 |
| Wore Gloves | 3,920 | 58 | 2,831 | 57 | 474 | 59.47 | 615 | 60 | 0.2137 |
| Wore Mask | 6,250 | 93 | 4,520 | 92 | 753 | 94.48 | 977 | 95 | <0.0001 |
| Stockpiled PPE | 1,948 | 29 | 1,536 | 31 | 169 | 21.2 | 243 | 24 | <0.0001 |
| Stockpiled Food | 2,659 | 39 | 2,089 | 42 | 268 | 33.63 | 302 | 29 | <0.0001 |
| Quarantined | 4,282 | 63 | 3,237 | 66 | 449 | 56.34 | 596 | 58 | <0.0001 |
| **Symptoms since last survey/in past month** |  |  |  |  |  |  |  |  |  |
| Covid-like symptoms (cough, fever, or shortness of breath) | 1,143 | 17 | 918 | 19 | 117 | 14.68 | 108 | 11 | <0.0001 |
| New Cough | 664 | 10 | 553 | 11 | 57 | 7.15 | 54 | 5 | <0.0001 |
| Fever | 312 | 5 | 260 | 5 | 33 | 4.14 | 19 | 2 | <0.0001 |
| Shortness Breath | 488 | 7 | 376 | 8 | 52 | 6.52 | 60 | 6 | 0.0984 |
| Coughing Blood | 52 | 1 | 50 | 1 | -- | -- | 2 | 0 | 0.0007 |

### COVID-19 symptoms and care outcomes

One in six (17% or N = 1,143) reported any COVID-like symptoms in the month prior to Visit 1 (cough, fever or shortness of breath) and this decreased significantly with age (19% versus 11% among 18-49 and 60+ year olds, respectively and p<0.001) (Table 3). The most common symptoms reported were new cough (10%) followed by shortness of breath (7%) and fever (5%). Among the 17% of participants reporting COVID-like symptoms (N = 1,143), 35% (N = 396) said they called or saw a physician/healthcare professional and 8% (N = 87) were hospitalized (Table 4). Compared to participants at lower risk for COVID-19 illness, participants with higher risk for severe COVID-19 illness were more likely to report seeing a physician or hospitalization (28% versus 38% and 2% versus 10%, respectively and p <0.001 for each comparison). Among all participants, 12% (N = 836) reported being tested for COVID-19 and 4% (N = 268) reported receiving a COVID-19 diagnosis. Participants at higher risk for COVID-19 illness were significantly more likely to report testing or receiving a diagnosis than participants at lower risk for severe COVID-19 illness (testing: 14% versus 10% and diagnosis: 5% versus 2%, respectively and p<0.001 for each comparison).

**Table 4:** CHASING COVID Cohort Care Outcomes Stratified by Risk for Severe COVID-19 Illness.

| Variable | Overall |  | No |  | Yes |  | Chi-Square<br>p-value |
| --- | --- | --- | --- | --- | --- | --- | --- |
|  | N | % | N | % | N | % |  |
| All with COVID-like symptoms: cough, fever, or shortness of breath | 1,143 | 100 | 366 | 32 | 777 | 68 |  |
| **Saw Physician** |  |  |  |  |  |  | **0.0042** |
| Yes | 396 | 35 | 102 | 28 | 294 | 38 |  |
| No | 741 | 65 | 262 | 72 | 479 | 62 |  |
| Missing | 6 | 1 | 2 | 1 | 4 | 1 |  |
| **Hospitalized** |  |  |  |  |  |  | **<0.0001** |
| Yes | 87 | 8 | 7 | 2 | 80 | 10 |  |
| No | 1,054 | 92 | 358 | 98 | 696 | 90 |  |
| Missing | 2 | 0 | 1 | 0 | 1 | 0 |  |
| All Participants | 6,753 | 100 | 2,989 | 100 | 3,764 | 100 |  |
| **Tested** |  |  |  |  |  |  | **<0.0001** |
| Yes | 836 | 12 | 291 | 10 | 545 | 14 |  |
| No, but tried to be tested | 743 | 11 | 290 | 10 | 453 | 12 |  |
| No, did not need to be tested | 5,134 | 76 | 2,389 | 80 | 2,745 | 73 |  |
| Missing | 40 | 1 | 19 | 1 | 21 | 1 |  |
| **COVID-19 Diagnosis** |  |  |  |  |  |  | **<0.0001** |
| Yes | 268 | 4 | 68 | 2 | 200 | 5 |  |
| No | 605 | 9 | 238 | 8 | 367 | 10 |  |
| Don't know | 26 | 0 | 12 | 0 | 14 | 0 |  |
| Missing | 5,854 | 87 | 2,671 | 89 | 3,183 | 85 |  |

### Baseline serologic testing

4,247 of 6,482 consenting participants (65.5%) provided a dried blood spot specimen for serologic testing, of whom 135 (3.2%, 95% CI 2.7%-3.5%) were positive by screening and 90 (2.1%, 95% CI 1.7%-2.6%) by confirmatory serologic testing. While differences were not statistically significant, seropositive persons by either test tended to be younger, male, and from the northeast U.S.

### Household factors and the risk of severe COVID-like illness early in the U.S. pandemic.^14^

Early in the U.S. pandemic, crowded indoor settings and sustained close indoor contact without masks were associated with an increased likelihood of SARS-CoV-2 spread. The role of such exposures, as well as the role of the presence of children in the household have not been investigated as risk factors for severe COVID-like illness (i.e., hospitalization). We found that the risk of hospitalization due to COVID-19 was higher among participants who had (versus those who did not have) children in the home, with an adjusted odds ratio [aOR] of hospitalization of 10.5 (95% CI 5.7-19.1) among study participants living in multi-unit dwellings and 2.2 (95% CI 1.2-6.5) among those living in single unit dwellings. Among participants living in multi-unit dwellings, the aOR for COVID-19 hospitalization among participants with more than 4 persons in their household (versus 1 person) was 2.5 (95% CI 1.0-6.1), and 0.8 (95% CI 0.15-4.1) among those living in single unit dwellings. This work demonstrated that, early in the U.S. SARS-CoV-2 pandemic, certain household exposures likely increased the risk of both SARS-CoV-2 acquisition and the risk of severe COVID-19 disease requiring hospitalization. These findings may have implications for mask wearing and other mitigation strategies at home during the initial phase of future ‘stay at home’ orders.

### The relationship between anxiety, health, and potential stressors among adults in the United States during the early phase of the COVID-19 pandemic.^15^

Epidemiologic data on the mental health impact of the COVID-19 pandemic in the U.S. remains limited. We found that more than one-third (35%) of individuals reported moderate or severe anxiety symptoms at cohort screening/enrollment visits. Having lost income due to COVID-19 (adjusted prevalence ratio [aPR] 1.27 (95% Confidence Interval [CI] 1.16, 1.30), having recent COVID-like symptoms (aPR 1.17 (95% CI 1.05, 1,31), and having been previously diagnosed with depression (aPR 1.49, (95% CI 1.35, 1.64) were positively associated with moderate or severe anxiety symptoms. This work demonstrated that anxiety symptoms were common and appear to be elevated among adults in the U.S. during the early phase of the COVID-19 pandemic. Strategies to screen and treat individuals at increased risk of anxiety, such as individuals experiencing financial hardship and individuals with prior diagnoses of depression, should be developed and implemented.

*SARS-CoV-2 Testing Service Preferences of Adults in the United States: Discrete Choice Experiment^16^ and Patterns of SARS-CoV-2 testing preferences in a national cohort in the United States.^17^* A discrete choice experiment (DCE) was used to assess the relative importance of type of SARS-CoV-2 test, specimen type, testing venue, and results turnaround time. Turnaround time for test results had the highest relative importance (30.4%), followed by test type (28.3%), specimen type (26.2%), and venue (15.0%). Participants preferred fast results on both past and current infection and using a noninvasive specimen, preferably collected at home. Simulations suggested that providing immediate or same day test results, providing both PCR and serology, or collecting oral specimens would substantially increase testing uptake over the current typical testing option. Using latent class analysis. we also found five distinct patterns in testing preferences in the cohort, including groups of ‘comprehensive testers’, ‘fast-track testers’, ‘dual testers’, ‘non-invasive, dual testers’, and ‘home testers’.^17^ We concluded that testing strategies that offer account for preferences and their heterogeneity would likely have the most uptake and engagement among residents in communities with increasing community transmission of SARS-CoV-2.

### Future plans

The CHASING COVID Cohort Study will continue to conduct ongoing monitoring of the cumulative incidence and determinants of SARS-CoV-2 outcomes (including mortality), mental health outcomes (e.g., anxiety and depression) and economic outcomes (e.g., income loss, food security), as well as overall and cause-specific mortality. A number of longitudinal analyses are ongoing or planned using the data that have already been collected, with priorities including those related to COVID-19 vaccine hesitancy, uptake and effectiveness; NPI engagement before and after vaccination; incidence, prevalence and correlates of long-haul COVID-19; extent and duration of protective effect of SARS-CoV-2 antibodies.

## DISCUSSION

This longitudinal cohort study enrolled 6,753 persons from all 50 U.S. states, the District of Columbia, Puerto Rico and Guam, and was rapidly established during the upswing of the SARS-CoV-2 pandemic in the U.S. The cohort is geographically and socio-demographically diverse, and includes participants from many active hotspots, as well as health care workers and other essential workers, and individuals who are vulnerable to severe outcomes associated with SARS-CoV-2 infection. Initial serologic testing indicates that the cohort overwhelmingly had no evidence of prior SARS-CoV-2 infection at the time of specimen collection, suggesting a high potential for observing subsequent seroconversions, and the extent of protective effects of new vaccines.

At cohort screening/enrollment, nearly one in six participants (17%) reported having had recent COVID-like symptoms. However, only small proportions of participants reported having been tested for or diagnosed with SARS-CoV-2 (12% and 4%, respectively). Participants with elevated risk for COVID-19 illness were more likely to report seeking care, hospitalization, and testing than participants without elevated risk. This is consistent with both the recommendations for testing and care seeking during the late spring and early summer of 2020, as well as the relative lack of SARS-CoV-2 testing availability in most parts of the U.S.

Recent evidence of waning of SARS-CoV-2 antibodies to both nucleocapsid and spike proteins, combined with the timing of specimen collection for many participants in our cohort, make our serology-based proxy measure for the cumulative incidence of SARS-CoV-2 infection challenging to interpret.^18^ Since SARS-CoV-2 antibodies may wane with time, and waning may be faster for antibodies to the nucleocapsid protein of the SARS-CoV-2 virus^19^, we have likely underestimated the cumulative incidence of infection in the cohort leading up to enrollment. Subsequent antibody testing in our cohort (S_2_) will help clarify whether participants who were positive on the screening test for total SARS-CoV-2 antibodies and negative on the follow-up IgG test represent early infections or false positives. However, antibody waning may also make this interpretation challenging, as up to 5 months may have passed between the first and second specimen collection for some participants. Recently, other teams have developed algorithms to correct seroprevalence estimates for waning^20^, and we may be able to use these approaches to correct our estimates of cumulative incidence in our cohort going forward. Limitations of serological assays notwithstanding^21^, recent cross-sectional serosurveys done prior to the relaxing of physical distancing have reported cumulative incidence estimates ranging from 1.7% in Indiana to 21% in New York City, and <10% nationally as of the end of September 2020.^18,22–26^

Strengths of our cohort study include its prospective design, allowing direct observation of seroconversions and incident COVID-19 disease among those who were unexposed and/or disease free at enrollment. We also have been able to include a range of relevant measures, for a more comprehensive view of the impact of the pandemic and its response on several outcomes in addition to SARS-CoV-2 infection, including mental health and economic outcomes, vaccine uptake, and long-haul COVID. The longitudinal design also allows prospective estimation of the incidence of COVID-19 disease among those with antibodies to SARS-CoV-2, allowing assessment of the extent to which SARS-CoV-2 antibodies, acquired through natural infection or through vaccination, offer short-term protection against subsequent disease. Prospective studies are complementary to, and offer some strengths over, cross sectional studies, especially in the context of rapidly evolving emergencies and the associated public health response. While repeat cross-sectional surveys are valuable in a pandemic, including their ability to assess trends in many important outcomes, they cannot assess what factors may influence change over time in an individual’s or a household’s exposures and outcomes.

Our study includes those who recover from SARS-CoV-2 infection (asymptomatic or mild) and COVID-19 disease, and follows these individuals over time to characterize their recovery, including persistence of symptoms, onset and persistence of new symptoms, and persistence of antibodies. This will allow us to characterize of the epidemiology of long-haul COVID (aka post-acute covid syndrome or PACS).^27^ We will also be able to assess mortality and related risk factors via matching with the US National Death Index.^28^

Our approach employs protocols for overcoming common pitfalls of fully online studies (e.g., repeat/duplicate participation). Our online, volunteer recruitment approach allows us to sample individuals who may not be reached by traditional telephone recruitment approaches, which can have very low response rates. As part of our enrollment procedures, we record IP address, email addresses, participant contact information, and require participants to have valid U.S. mailing addresses (required for those who opt to receive an at-home SARS-CoV-2 specimen collection kit). Participants are “known” to the research team (name, email, address), thus averting some of the traditional shortcomings of online-only studies (particularly anonymous, cross-sectional online studies).

Our cohort study has limitations worth noting, as they inform what can and cannot be assessed. First, while our study is national in scope, we will be unable to provide estimates of seroprevalence and cumulative incidence that are representative of that of the U.S. population. Second, we will likely underestimate the incidence of COVID-19 related hospitalizations. Most research studies deployed in the middle of a pandemic, including ours, will by definition, produce some biased estimates since they will not include some information on participants who died from COVID-19, were hospitalized with COVID-19 after recruitment and were either lost to follow-up or were too sick to participate in the research activities. We will ascertain deaths in our participants using the National Death Index. Additionally, from published studies, we will routinely assess bias in our estimates due to these factors and adjust them accordingly when possible. Third, we will largely be unable to conduct state or county specific analyses, except for a few localities with high participation (e.g., New York, Texas, Florida and California). Finally, although our sample is not representative of the entire U.S. population, it is geographically representative and socio-demographically diverse. Our study will complement other efforts and approaches to address similar research questions, such as the NIH’s national study^29^, and the COVID-Vu study.^30^ Indeed, it will be important to assess the extent to which the different strategies used in each of these cohort studies reach similar or divergent conclusions.

### Collaboration, data sharing, and dissemination of findings

We will post a deidentified, HIPAA compliant, public use version of Visit 1 and follow-up data on GitHub.^31^ Data will be presented as flat text files (CSV) formatted for compatibility with county-level longitudinal case load datasets, including date, county, state, and fips code. We will continue to provide direct feedback to our cohort and other stakeholders who have signed up for updates via follow-up emails to participants, and the CUNY ISPH study website.^32^

## Conclusion

A geographically and socio-demographically diverse group of participants, largely without serologic evidence of prior SARS-CoV-2 infection, was rapidly enrolled into a national longitudinal cohort study during the upswing of the SARS-CoV-2 pandemic in the U.S. Strengths of the study include the potential for direct observation of seroconversions and incident COVID-19 disease in areas of active transmission, and related determinants. The study is also examining a range of other outcomes, including psychosocial and employment outcomes, and long-haul COVID. The study has the potential to monitor and assess the uptake and impact of the public health response to control and mitigate the SARS-CoV-2 pandemic in the U.S., including the uptake of recently authorized vaccines.

## Data Availability

The data that support the findings of this study are available on request from the corresponding author, [DN]. The data are not yet publicly available, but we are preparing to post a deidentified, HIPAA compliant, public use version of our baseline and follow-up data on GitHub.

## Contributors

MMR, SGK, CG and DN conceptualized the study. SK, MC and MMR performed statistical analyses. MR and DN wrote the first draft of the paper. MMR, SGK, AB, CM, SK, MR, WY, AM, RZ, DW, CG, AP, LW, and DN contributed to interpreting the data and to the writing and revising of the manuscript.

## Acknowledgements

The authors wish to thank the participants of the CHASING COVID Cohort Study. We are grateful to you for your contributions to the advancement of science around the SARS-CoV-2 pandemic.

